# A novel assessment method for COVID-19 humoral immunity duration using serial measurements in naturally infected and vaccinated subjects

**DOI:** 10.1101/2021.12.28.21268183

**Authors:** Jasper de Boer, Ursula Saade, Elodie Granjon, Sophie Trouillet-Assant, Carla Saade, Hans Pottel, Maan Zrein, on behalf of Covid ser study group

## Abstract

**Background:** It is crucial for medical decision-making and vaccination strategies to collect information on sustainability of immune responses after infection or vaccination, and how long-lasting antibodies against SARS-COV-2 could provide a humoral and protective immunity, preventing reinfection with SARS-CoV-2 or its variants. The aim of this study is to present a novel method to quantitatively measure and monitor the diversity of SARS-CoV-2 specific antibody profiles over time.

**Methods:** Two collections of serum samples were used in this study: A collection from 20 naturally infected subjects (follow-ups to 1 year) and a collection from 83 subjects vaccinated with one or two doses of Pfizer BioNtech vaccine (BNT162b2/BNT162b2) (follow-ups to 6 months). The Multi-SARS-CoV-2 assay, a multiparameter serology test, developed for the serological confirmation of past-infections was used to determine the reactivity of six different SARS-CoV-2 antigens. For each patient sample, 3 dilutions (1/50, 1/400 and 1/3200) were defined as an optimal set over the six antigens and their respective linear ranges, allowing accurate quantitation of the corresponding six specific antibodies. Nonlinear mixed-effects modelling was applied to convert intensity readings from 3 determined dilutions to a single quantification value for each antibody.

**Results:** Median half-life for the 20 naturally infected *vs* 74 vaccinated subjects (two doses) was respectively 120 *vs* 50 days for RBD, 127 *vs* 53 days for S1 and 187 *vs* 86 days for S2 antibodies. Respectively, 90% of the antibody concentration wanes after 398 *vs* 158 days for RBD, 420 *vs* 171 days for S1, and 620 *vs* 225 days for S2 after the second vaccine shot.

**Conclusion:** The newly proposed method, based on a series of a limited number of dilutions, can convert a conventional qualitative assay into a quantitative assay. This conversion helps define the sustainability of specific immune responses against each relevant viral antigen and can help in defining the protection characteristics after an infection or a vaccination.

## Introduction

The Severe acute respiratory syndrome coronavirus 2 (SARS-CoV-2) was first described in December 2019 (1) and has rapidly spread and continues to cause a major health crisis around the world since then. The virus causes the respiratory illness COVID-19, and the outbreak was declared a pandemic on 11 March 2020 by the WHO (2). The COVID-19 pandemic represents the greatest medical and socio-economic challenge of our time. As there is no antiviral drug to treat the disease at this moment, it is crucial for medical decision-making and vaccination strategies to collect information on sustainability of immune response after infection and define for how long antibodies against SARS-COV-2 could provide a humoral immunity, preventing reinfection with SARS-COV-2 or its variants. Now that massive vaccination programs have been implemented, it is equally important to understand the duration of vaccine-induced immunity and anticipate possible modifications of the vaccination strategies. At this moment, a consistent depiction of the humoral immune response after natural SARS-COV-2 infection is not available.

So far, studies suggest that antibody levels may decrease rapidly in infected individuals, and this mainly depends on the severity of the infection (asymptomatic, mildly symptomatic, or severely symptomatic requiring hospitalization and referral to the ICU). (3), (4), (5)

The duration of immunity is, of course, a very important aspect for protection following the natural infection or after vaccination. Beside cellular immunity, virus-specific antibodies represent an important component for protection and large efforts are being made globally to set objective criteria to define at individual level the duration of such protective immunity. (6), (7), (8), (9)

Long et al. (10) described a decline in antibody titers in the convalescent phase of the disease, suggesting that antibodies to SARS-CoV-2 may fade away quickly. Furthermore, Prévost et al. (11) evaluated 98 infected patients and found that while most individuals developed neutralizing antibodies within the first two weeks of infection, the level of neutralizing activity decreased significantly over time. Therefore, measuring the kinetics of binding- and neutralizing-antibodies remains to be explored, preferably using a simple and universal method that can be implemented in a wide majority of moderate-resources laboratories.

In this study we present a novel method to measure and monitor SARS-CoV-2 specific antibody profiles over time, using the Multi-SARS-CoV-2, a microplate-based immunoassay (InfYnity Biomarkers, Lyon, France), that can detect the presence of 6 different SARS-CoV-2 specific antibodies in a single well of a 96-well microplate thus reducing inter-assay variability. Unlike conventional immunoassay methods that provides binary results, this approach provides serological patterns with a high level of relevant information.

## Methods

### Study design, population, and origin of the samples

Two collections of serum samples were used in this study: A collection from naturally infected subjects and a collection from vaccinated subjects. The first collection consisted of 101 serum samples and originated from 20 naturally infected patients out of which 17 were followed up over time, up to 360 days (median=268). For each patient two to nine longitudinal samples were available (median=5). Patients were all symptomatic, mainly with fever, coughing, having shortness of breath, muscle pain and general weakness. Most patients were confined at home with ambulatory treatment, only a few were hospitalized. Serum samples from these 20 patients were collected for up to 360 days and measured with the Multi-SARS-CoV-2 immunoassay to demonstrate the monitoring capabilities of the new assay and to investigate the antibody dynamics of naturally infected patients.

The second collection is a prospective longitudinal cohort study conducted at the laboratory associated with the National reference center for respiratory viruses (University Hospital of Lyon, France). Eighty-three (HCW) vaccinated with one or 2 doses of Pfizer BioNtech vaccine (n=83; BNT162b2/BNT162b2) were included. Blood samples were collected i) before the first dose of vaccine, ii) before the second injection of vaccine corresponding to 4 weeks after the first dose for participants, iii) 4 weeks and six months after the full vaccination. The pre-vaccination blood sample was only used to document a previous SARS-CoV-2 infection. Participants who have been previously infected with SARS-CoV-2 (convalescent group, 11%) have only one injection, therefore the second sample was omitted. One participant was infected with SARS-CoV-2 between the 2 doses and therefore got a single dose of the vaccine. Serological investigation with commercial assays was performed on the 323 samples.

For each naturally infected patient, 3 dilutions (1/50, 1/400 and 1/3200) at each time-point were assayed, while 4 dilutions (1/50, 1/100, 1/400 and 1/3200) were available at each time-point for the vaccinated subjects.

### Ethical Statements

For the vaccinated subjects, a written informed consent was obtained from all participants; ethics approval was obtained from the national review board for biomedical research in April 2020 (Comité de Protection des Personnes Sud Méditerranée I, Marseille, France; ID RCB 2020-A00932-37), and the study was registered on ClinicalTrials.gov (NCT04341142).

For the naturally infected covid patients, immune serum samples collected in 2020 and 2021 were acquired from ABO Pharmaceuticals (ABO Pharmaceuticals, SanDiego, CA, USA) a duly authorized blood banking organization.

### Multi-SARS-CoV-2 assay

The Multi-SARS-CoV-2 immunoassay was used to monitor detailed serological profiles. The multiplex technology allows multiple parameters to be combined in a single well of a 96-well microplate. The printing process is based on non-contact piezo electric impulsion of a defined volume of an antigenic solution. The bioprinting was performed to print six different SARS-CoV-2 specific antigens: 1) MP1; specific epitope for SARS-CoV-2 membrane antigen; enhanced according to disease severity. 2) NP1; full-length nucleocapsid recombinant protein antigen. 3) NP2; specific epitope of the nucleocapsid protein, enhanced according to disease severity. 4) RBD; recombinant Receptor Binding Domain of the spike protein. 5) S1; recombinant spike protein S1. 6) S2; recombinant spike protein S2.

Antigens were printed at the bottom of each well at precise X-Y coordinates, under controlled humidity and temperature conditions. Each antigen spot is printed in duplicate to improve fault tolerance. Positive control spots were printed in quadruplicate to define a precise spatial orientation pattern and validate the correct sequential distribution of all biological and chemical materials as well as operating performance (human serum samples, enzyme conjugate, and substrate).

Prior to this study, assay validation was conducted to ensure that all performance attributes are in line with In-vitro Diagnostic and quality requirements. In brief, each antigen is analyzed for its intrinsic specificity and sensitivity using a set of seropositive (n=540; all >14 days post-PCR determination) and seronegative samples (n=270; including pre-pandemic samples) and optimal concentration for each antigen is determined by Receiver Operating Characteristic curves (ROC).

For performance evaluation the NIBSC calibrator was used (ref. 20/136) to define the lower limit of detection for each antibody specificity.

Each plate was read and analyzed using a colorimetric image analyzer. The software calculates the median pixel intensity for each spot with the background noise subtracted. To establish the net intensity for each antigen, the mean value of duplicated spots was calculated. Mean spot intensities for each antigen were normalized (or scaled) by dividing the value by the average positive control intensity, which show the maximum attainable intensity in the assay, with the aim to reduce assay variability and to have a common value range of [0, 100] for all tested antigens.

### Dilution sequence

Biomarker reactivity curves that express the relationship between log antibody concentration and intensity typically show a sigmoidal shape that levels off at both ends of the curve. Consequently, at high intensities, a small change in intensity leads to a considerable change in antibody concentration. Since minor fluctuations may always be expected due to test variability, the nonlinear plateau-range is ill-suited for an accurate quantification. The Multi-SARS-CoV-2 assay was developed for the serological confirmation of past infections and was therefore optimized to produce strong signals with sera from infected patients and show no or non-significant signals with sera from uninfected persons. Readings in the plateau-range can therefore be expected but make quantitation, and therefore monitoring of antibodies dynamic difficult.

To ensure a reading of each antigen in the linear range, samples from the longitudinal sets were serially diluted. The dilution factors were chosen in such a way that overlap of linear ranges for all antigens was assured, observed maximal concentrations could be quantified and the number of dilutions was minimized (to reduce the operational cost). The optimal set of dilutions and their corresponding dilution factors were determined *a-priori* by testing a subset of the longitudinal collection. Forty samples were tested, 2 samples per patient, using a dense dilution sequence with a step of factor two (1/50, 1/100, 1/200, 1/400, 1/800, 1/1600, 1/3200 and 1/6400). Finally, we selected the set of 3 dilutions (1/50, 1/400 and 1/3200) as an optimal set over the 6 antigens and their respective linear ranges, allowing accurate quantitation of the corresponding 6 specific antibodies. The dilution results are converted into a quantitative antibody value using statistical modelling.

## Statistical methods

Nonlinear mixed-effects modelling was applied to convert intensity readings from the 3 (or 4) determined dilutions to a single quantification per antibody. The used model is based on the standard 2-parameter logistic curve (sigmoidal standard curve with fixed top = 100 and bottom = 0) and is described in the following Equation 1.

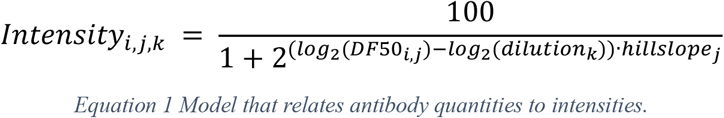

For both collections combined, intensities were available for I=396 samples (coming from 104 subjects at different time-points), J=7 antibodies and K=3 or 4 dilutions. In the above equation, *Intensity*_*i,j,k*_ corresponds to the intensity of the i^th^ sample, j^th^ antibody and k^th^ dilution. The Dilution Factor 50 (DF50) value corresponds to the estimated dilution at which an intensity value of 50% is observed. This value summarizes the 3 (or 4) antibody intensities and represents a quantitative information of antibody concentration. Although the unit of DF50 is arbitrary, its relationship with concentration is linear. The R ‘LME4’ package was utilized for model fitting. (12) The model fit results into 6 (antibody) DF50 values for each individual patients’ time-point.

As an example, Figure 1 shows the RBD fits for 3 time-points of one naturally infected subject (subject #6). At dilution 1/50 the 29 days, 95 days and 270 days serum samples show high and saturated optical densities (or biomarker reactivities) and are thus unquantifiable without the dilution sequence. The fitted sigmoid curves on the three dilutions shift position over time. The position of the curve is summarized by the DF50-value, that is, the estimated dilution factor corresponding to 50% reactivity. In figure 1, the DF50s are located at the points where the dashed line at 50% crosses the curves. In the above-described example, the complete curve clearly shifts with time (corresponding to higher dilution factor). Note that the location of the sigmoid curve and thus the DF50 value is obtained from the fitted curve for each sample (thus for each subject at each time-point), based on the measured ODs of the 3 tested dilutions. The hillslopes (per marker) are, however, fitted on the combination of all 396 samples. This example clearly illustrates the need to use the dilution approach for quantitative measurements. It is also possible to convert the 2-parameter sigmoidal function to a linear equation, allowing to determine hillslope and DF50 for each individual sample (see Supplement). Extrapolation beyond DF = 3200 was considered acceptable when not exceeding DF = 6400 (when DF was greater than 6400 it was fixed to 6400).

**Figure 1.**
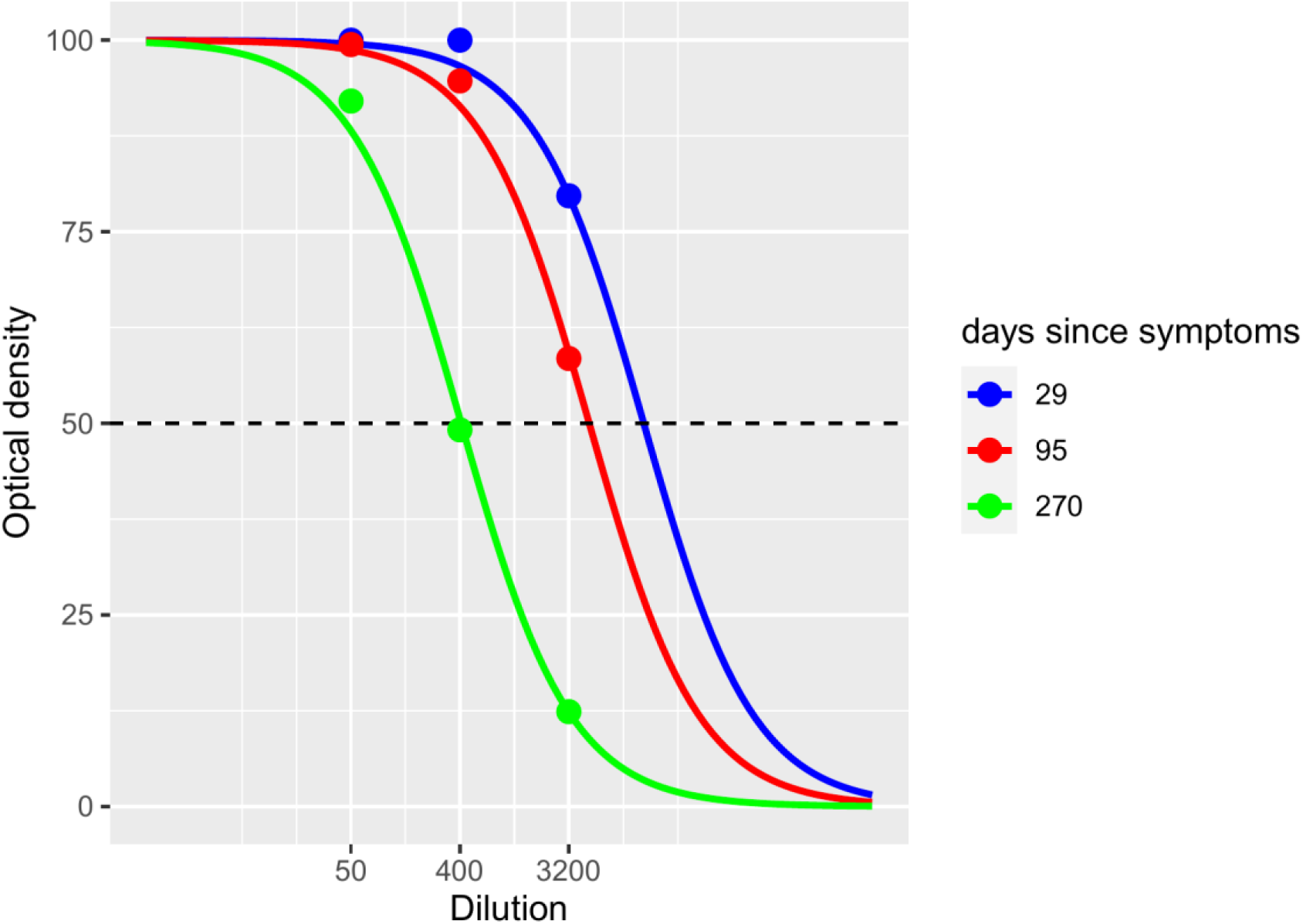
Sigmoidal curve fitting of 3 dilutions (1/50, 1/400, 1/3200) for 3 timepoints of the naturally infected subject #6 for the anti-RBD antibodies.

## Results

Patient characteristics of the naturally infected subjects are shown in Table 1.

**Table 1.** Patient characteristics

| Characteristic | Naturally infected cohort | Vaccinated cohort |
| --- | --- | --- |
| **Sex (Male/Female)** | 60%/40% (12/8) | 81% / 19% (67/16) |
| **Age (Years)** | 43 ± 10 (range: 23 – 60) | 47 ± 11 (range: 22 – 76) |
| **Median # samples** | 5 (IQR = 2; range: 2 – 9) | 3 or 4 |
| **Follow-up time (days)** | 268 (IQR = 89; range: 127 – 360) | 180 |

Figure 2 shows the evolution of each (log-transformed) antigen concentration for the 20 patients and 6 markers of the naturally infected subject’s collection. The general time-evolution trend displays a log-linear decline of concentration, which supports constant half-life for at least the duration of the study period. The graphs also show that decline rates differ between patients and antigens. A linear regression model was used to fit the log-linear decline per patient and per marker to model the individual dynamics (see Supplement). The results are shown in figure 2 and summarized in Table 2 for the naturally infected subjects. The DF50-values for the first sample of each patient and the calculated half-life are listed in the Supplement (Table S1).

**Figure 2.**
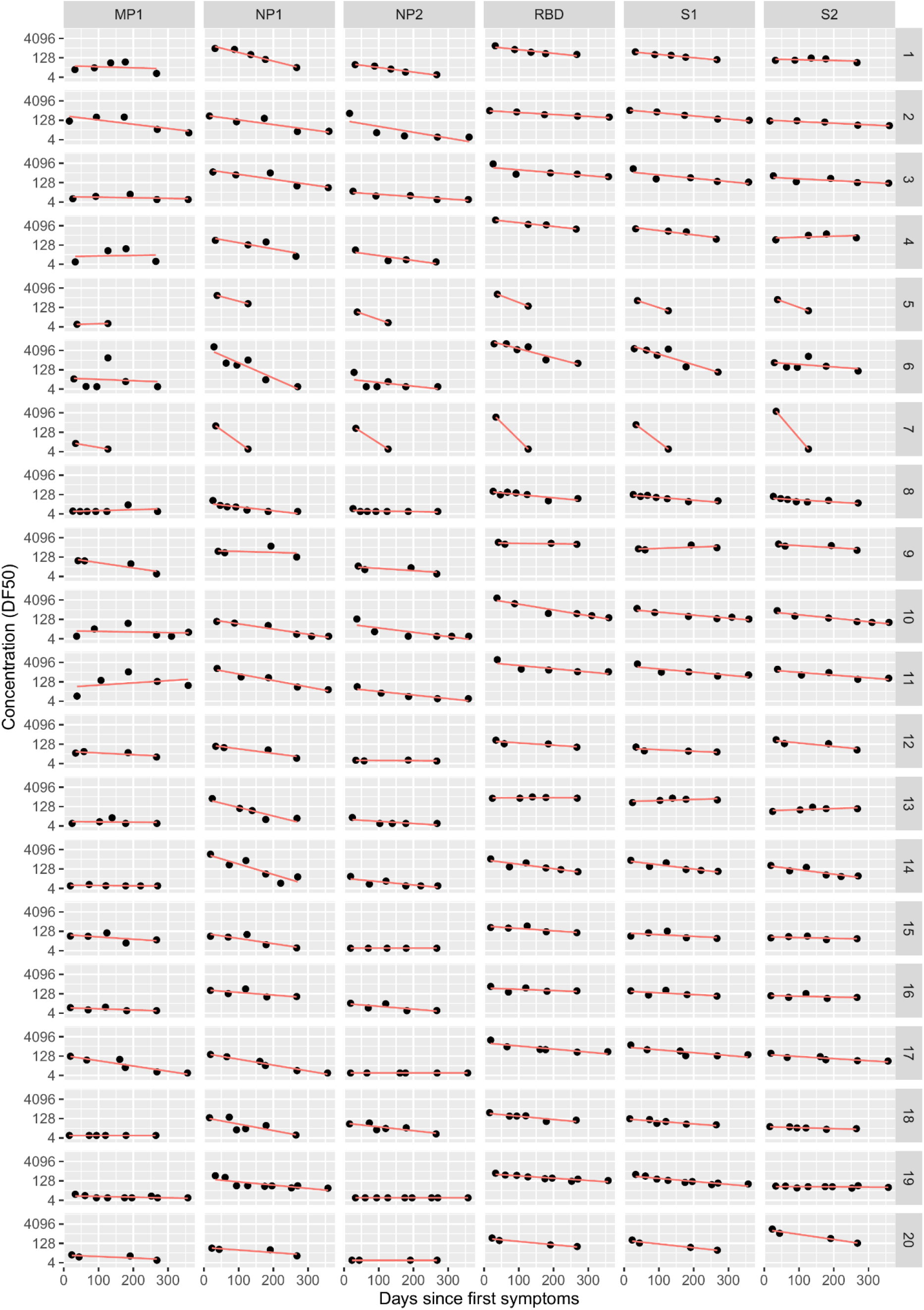
Antibody concentrations over time for 20 naturally infected patients.

**Table 2.**
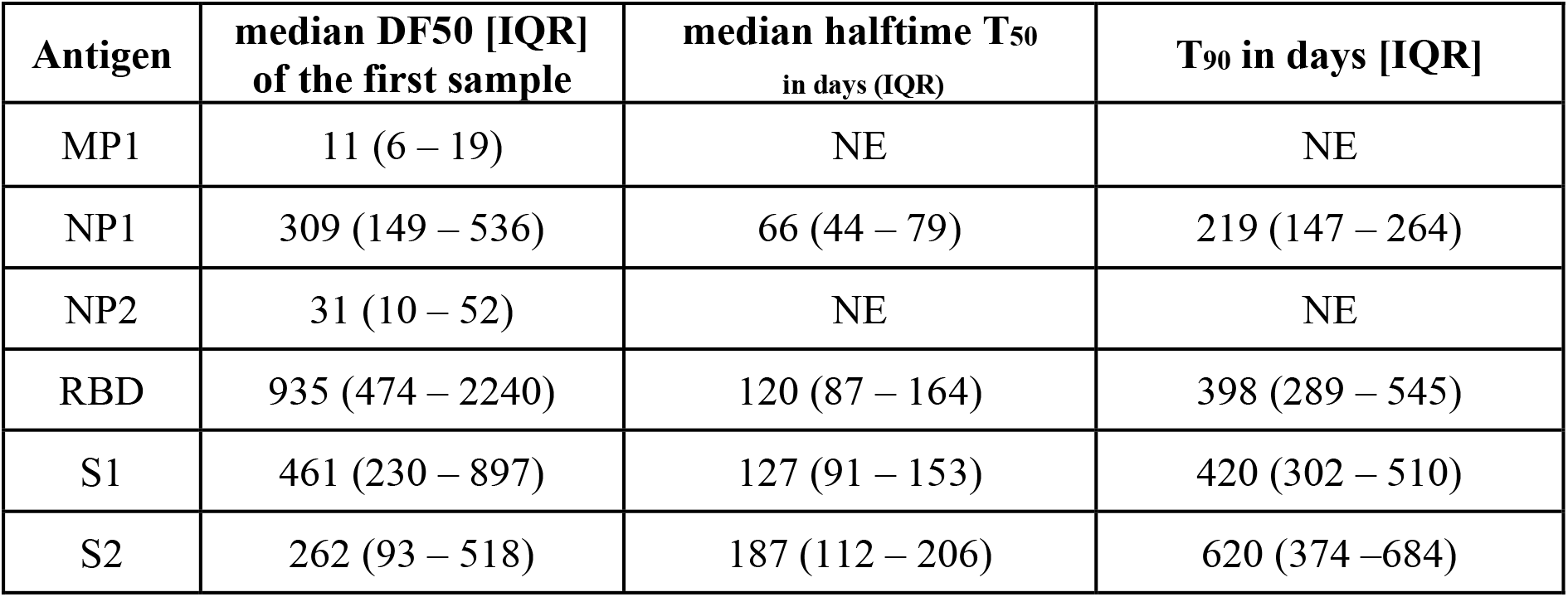
Summary of the decline dynamics for each antigen for the 20 naturally infected subjects. DF50 < 50 of the first sample corresponds to no or little reactivity. NE = Not Estimable because of low reactivity.

As for the vaccinated individuals, the median DF50-values at the different time-points (T0, T1, T2 and T3) are presented in Table 3. There were 74 subjects who were vaccinated twice as they (self-)reported no prior COVID infection, and 10 subjects with a self-reported prior COVID infection who were only vaccinated once (no data at time-point T1). However, among the 74 subjects who were vaccinated twice, we identified 6 subjects who showed antigen reactivity at baseline (T0). The characteristic times T_50_ and T_90_, obtained from the DF50-values at the two time-points T2 and T3 are summarized in Table 4. These characteristic times are estimated from 2 points only, assuming an exponential decay. As this might be prone to error, we also calculated the relative loss in DF50-reactivity between T2 and T3 (assuming a linear decay between these two time-points). Taking RBD as the reference, there is a loss of about 90% in antibodies at time T3, which is 6 months after the second vaccine shot.

**Table 3.** Summary of the dynamics for each antigen for the 83 vaccinated subjects. Median DF50-values [IQR] for RBD, S1 and S2 are presented at the different time-points T0 (baseline), T1 (3 weeks after first vaccine), T2 (1 month after second vaccine) and T3 (6 months after second vaccine) for group 1 (n = 73) vaccinated subjects who had no prior COVID infection, for group 2 (n = 9) vaccinated subjects who had a prior COVID infection and for group 3 (n = 6) vaccinated subjects who reported no prior COVID infection, but who showed antibodies reactivity at baseline (T0). The other antibodies (MP1, NP1 and NP2) were not reported as they were not triggered by the vaccines.

| Group | Antigen | T0 | T1 | T2 | T3 |
| --- | --- | --- | --- | --- | --- |
| 1<br>(n = 74) | RBD | ≤6 [≤6 – 8] | 261 [122 – 551] | 3123 [2225 – 7076] | 460 [277 – 735] |
|  | S1 | ≤6 [≤6 – ≤6] | 104 [53 – 205] | 1530 [873 – 2727] | 245 [140 – 453] |
|  | S2 | ≤6 [≤6 – 12] | 32[18– 56] | 109 [51 – 193] | 29 [20 – 53] |
| 2<br>(n = 9) | RBD | 148 [42 – 257] | - | 6448 [5650 – 10316] | 1473 [1264 – 716] |
|  | S1 | 60 [20 – 66] | - | 4773 [3384 – 5488] | 833 [716 – 865] |
|  | S2 | 45 [27 – 62] | - | 1501 [936 – 1626] | 126 [74 – 164] |
| 3<br>(n = 6) | RBD | 38 [22 – 67] | 3121 [144– 5936] | 4018 [3339– 8527] | 1211 [527– 2910] |
|  | S1 | 15 [8 – 33] | 1802 [56– 4317] | 2278 [1466– 7109] | 708 [375– 1890] |
|  | S2 | 21 [12 – 42] | 388 [39– 1283] | 327 [118– 1028] | 108 [57– 208] |

**Table 4.** Characteristic times for the waning antibodies. The half-life T_50_ (T_90_) equals the number of days at which there is 50% (10%) left from the estimated DF50 at time zero assuming an exponential decay. The estimated % relative loss is calculated as [DF50(T2) - DF50(T3)] / DF50(T2). We compared specific antibodies between groups 1 and 2 using a Wilcoxon rank sum test: RBD, p = 0.330; S1, p = 0.855: S2, p = 0.001.

| Group | Antigen | T<sub>50</sub> in days (IQR) | T<sub>90</sub> in days [IQR] | Estimated % relative loss between T2 and T3 |
| --- | --- | --- | --- | --- |
| 1<br>(n = 74) | RBD | 50 [42 – 58] | 168 [140 – 193] | 87% [82 – 91] |
|  | S1 | 53 [45 – 68] | 177 [149 – 227] | 86% [77 – 90] |
|  | S2 | 86 [59 – 123] | 287 [197 – 309] | 69% [56 – 81] |
| 2<br>(n = 9) | RBD | 55 [48 – 61] | 182 [160 – 203] | 84% [76 – 86] |
|  | S1 | 49 [46 – 65] | 164 [153 – 214] | 84% [71 – 89] |
|  | S2 | 37 [37 – 51] | 122 [121 – 168] | 92% [71 – 94] |
| 3<br>(n = 6) | RBD | 84 [62 – 102] | 278 [205 – 340] | 0.71 [0.64 – 0.82] |
|  | S1 | 82 [78 – 91] | 272 [258 – 302] | 0.72 [0.68 – 0.74] |
|  | S2 | 122 [94 – 155] | 404 [312 – 514] | 0.58 [0.49 – 0.67] |

In addition, we compared median half-lives for the 20 naturally infected *vs* 74 vaccinated subjects (two doses) using a Wilcoxon rank sum test. Median half-lives were respectively 120 *vs* 50 days for RBD (p < 0.001), 127 *vs* 53 days for S1 (p < 0.001), and 187 *vs* 86 days for S2 (p < 0.001) antibodies.

## Discussion

Quantitative measurement of humoral immune response provides an easy and robust surrogate marker of protection. In this study we present a novel method to quantify antibodies after SARS-CoV-2 infection over time, allowing us to predict the half-life of the humoral immune response. Determining the duration of the presence of antibodies is an important step towards determination of the duration of protective humoral immunity. If a natural infection induces a sustained and protective immunity, or at least, a long-lasting protection, it may enable the establishment of collective herd immunity. In addition to cellular immunity, antibodies are valuable surrogate indicators of protection against reinfection. Therefore, accurate determination of the duration of the presence of antibodies is essential. Based on the novel multiparametric method presented in this study, we were able to determine the half-life of antibodies for different antigens of naturally infected patients with median values of 120 days for RBD, 127 days for S1 and 187 days for S2, since the first signs of the infection, but with large individual variability. It takes about 1 year (398 days) to lose approximately 90% of the RBD reactivity. It should also be noted that the 20 patients in this study all showed mild to moderate symptoms. The time course as well as the duration of humoral immunity responses may well be entirely different following asymptomatic or severe infections. Patients who experience high viral replication during COVID-19 infections, typically express severe clinical outcomes and show high levels of humoral immunity. However, waning antibodies operate in a stealth mode and may not be captured by conventional immunoassays. Such waning can more easily be perceived when monitoring each individual antibody specificity instead of collectively measuring a global and consolidated immune response. The method we describe in this manuscript may serve as a tool to estimate the half-life of each antibody response as an attempt for establishing criteria of protection and for susceptibility to reinfection.

The same information is critical after vaccination. Indeed, there is no guarantee that a vaccine will give long lasting protection. The same method, described here, can easily be applied after vaccination to determine the half-life of the different antibodies. It may be particularly important for RBD and the S-protein since these titers correlate with neutralizing activity and are typically associated with early virus control. From our analysis, it should be emphasized that the time course as well as the duration of humoral responses after vaccination are entirely different compared to natural (asymptomatic, mildly symptomatic, or severely symptomatic) infections. Half-lives were significantly shorter with only 50 days for RBD, and it takes approximately only 6 months to lose 90% of the antibodies in subjects who received two vaccine shots (compared to about 1 year after a natural infection).

The role of the individual antibodies measured in the Multi-SARS-CoV-2 assay still has to be investigated. While anti-RBD and anti-S antibodies may be related to neutralization activity, the role of the membrane protein antigen (MP1) and the nucleocapsid recombinant protein antigens (NP1 and NP2) still need to be determined. When comparing the different antibodies’ concentration values of naturally infected subjects with vaccinated subjects a clear difference in NP1 and NP2 concentrations was observed. This suggests that the Multi-SARS-CoV-2 assay has the ability to distinguish the immune response of naturally infected and vaccinated subjects.

The half-life predicted by the nucleocapsid recombinant protein antigen NP1 was only about half of the estimated half-life of RBD, S1, S2. In vaccinated subjects who had a prior COVID infection, baseline values of NP1 and NP2 were similar (that is, quite low) to those of the baseline values of subjects who did not declare a prior COVID infection, suggesting that these antibodies disappear much faster after infection than the other antibodies.

Several studies reported significant reduction of humoral response to SARS-CoV-2 within 4 months after the diagnosis of COVID-19 (13), (4), corresponding to the half-life for RBD (116 days) and S1 (113 days) reported here.

Using sequential serum samples collected up to 94 days post onset of symptoms (POS) from 65 RT-qPCR confirmed SARS-CoV-2-infected individuals, showed sero-conversion in > 95% of cases and neutralizing Antibody (nAb) responses when sampled beyond 8 days POS. The authors observed that the magnitude of the nAb response is dependent upon disease severity, but this was not found to affect the kinetics of the nAb response. Declining nAb titers were observed during the follow-up period. Whilst some individuals with high peak response (>10 000) maintained titers >1000 at >60 days POS, some with lower peak had titers approaching baseline within the follow up period. A similar decline in nAb titers was also observed in a cohort of seropositive healthcare workers from 2 different hospitals. (14)

Iyer et al. measured the kinetics of early antibody responses to the RBD of the S protein of SARS-CoV-2 in a cohort of 259 symptomatic North American patients (up to 75 days after symptom onset) compared to antibody levels in 1548 individuals whose blood samples were obtained prior to the pandemic. IgG antibodies against RBD lasted longer and persisted through 75 days post-symptoms. IgG antibodies to SARS-CoV-2 RBD were highly correlated with nAb targeting the S protein. (15)

Studies of Dan et al. and Wu et al. have provided evidence that the circulating immune memory to SARS-CoV-2 appears to endure for more than 5 months in patients with previous SARS-CoV-2 infection. (16), (17)

Our study confirms these last findings. Indeed, despite the heterogeneity of immune responses, our results show that durable immunity against secondary COVID-19 disease is a possibility for most individuals. Our limited data indicate sustained humoral immunity in recovered patients who had symptomatic COVID-19, suggesting prolonged immunity. The half-lives reported in our study (about 120 days) are consistent with those reported by Dan et al., who mentioned that immunoglobulins (Ig)G targeting the SARS-CoV-2 spike protein were found relatively stable over time (half-life, 140 days; 95% CI, 20–240 days), with SARS-CoV-2 spike-specific memory B cells being even more abundant six months after SARS-CoV-2 infection than in earlier period. (17)

In vaccinated subjects, antibodies targeting MP1, NP1 and NP2 were always low, in line with the nature of the BNT162b2/Comirnaty vaccine that doesn’t induce such antibodies and solely expresses spike antigen inducing RBD, S1 and S2 antibodies. These antibody activations by the vaccines result in much higher reactivities compared to their corresponding concentrations in naturally infected patients, but this did not result in longer half-life showing that the velocity of antibody waning does not necessarily depend on the initial concentrations, but rather on the difference between a natural infection and vaccination. Anti-NP1 and anti-NP2 antibodies levels are correlated with disease severity. Naturally infected patients in our study did not experience severe infections, mostly with mild symptoms. Also, anti-NP1 and anti-NP2 antibodies had shorter half-life compared to the other antibodies, suggesting a faster decline. Vaccinated patients with prior a COVID infection were health-care workers with mild COVID symptoms, and the duration between the pre-infection timepoint and the post-vaccination timepoint was approximately a few months, indicating that the levels of antibodies against NP1 and NP2 had disappeared at the time of vaccination.

In conclusion, the newly proposed method, based on a series of a limited number of dilutions, can convert a conventional assay from qualitative testing into a quantitative assay, allowing to quantify the time decay of antibody response and to calculate half-life for these antibodies. This new procedure allows to collect information on sustainability of immune response, helping to understand and estimate the duration of humoral immunity, after infection or vaccination.

## Data Availability

All data produced in the present study are available upon request to the authors

## Notes

### Competing Interest Statement

Maan Zrein acts as Chief Scientific Officer employed by Infynity Biomarkers
Ursula Saade is employed at Infynity Biomarkers
Elodie Granjon was employed at Infynity Biomarkers

### Clinical Trial

NCT04341142

### Funding Statement

This study did not receive any funding

### Author Declarations

For the vaccinated subjects, a written informed consent was obtained from all participants; ethics approval was obtained from the national review board for biomedical research in April 2020 (COMITE DE PROTECTION DES PERSONNES SUD MEDITERRANEE I, Marseille, France; ID RCB 2020-A00932-37), and the study was registered on ClinicalTrials.gov (NCT04341142). For the naturally-infected COVID patients, immune serum samples collected in 2020 and 2021 were acquired from ABO Pharmaceuticals (ABO Pharmaceuticals, SanDiego, CA, USA) a duly authorized blood banking organization.

## References

1. Zhu N, Zhang D, Wang W, Li X, Yang B, Song J, et al. A Novel Coronavirus from Patients with Pneumonia in China, 2019. N Engl J Med. 2020 Feb 20;382(8):727–33.

2. WHO Director-General’s opening remarks at the media briefing on COVID-19 - 11 March 2020 [Internet]. [cited 2021 Dec 8]. Available from: https://www.who.int/director-general/speeches/detail/who-director-general-s-opening-remarks-at-the-media-briefing-on-covid-19 11-march-2020

3. Long Q-X, Tang X-J, Shi Q-L, Li Q, Deng H-J, Yuan J, et al. Clinical and immunological assessment of asymptomatic SARS-CoV-2 infections. Nat Med. 2020 Aug;26(8):1200–4.

4. Ibarrondo FJ, Fulcher JA, Goodman-Meza D, Elliott J, Hofmann C, Hausner MA, et al. Rapid Decay of Anti–SARS-CoV-2 Antibodies in Persons with Mild Covid-19. N Engl J Med. 2020 Sep 10;383(11):1085–7.

5. Liu L, To KK-W, Chan K-H, Wong Y-C, Zhou R, Kwan K-Y, et al. High neutralizing antibody titer in intensive care unit patients with COVID-19. Emerging Microbes & Infections. 2020 Jan 1;9(1):1664–70.

6. Perreault J, Tremblay T, Fournier M-J, Drouin M, Beaudoin-Bussières G, Prévost J, et al. Waning of SARS-CoV-2 RBD antibodies in longitudinal convalescent serum samples within 4 months after symptom onset. Blood. 2020 Nov 26;136(22):2588–91.

7. Wajnberg A, Amanat F, Firpo A, Altman DR, Bailey MJ, Mansour M, et al. Robust neutralizing antibodies to SARS-CoV-2 infection persist for months. Science. 2020 Dec 4;370(6521):1227–30.

8. Choe PG, Kim K-H, Kang CK, Suh HJ, Kang E, Lee SY, et al. Antibody Responses 8 Months after Asymptomatic or Mild SARS-CoV-2 Infection. Emerg Infect Dis. 2021 Mar;27(3):928–31.

9. Khoury J, Najjar-Debbiny R, Hanna A, Jabbour A, Abu Ahmad Y, Saffuri A, et al. COVID-19 vaccine – Long term immune decline and breakthrough infections. Vaccine. 2021 Nov;39(48):6984–9.

10. Long Q-X, Tang X-J, Shi Q-L, Li Q, Deng H-J, Yuan J, et al. Clinical and immunological assessment of asymptomatic SARS-CoV-2 infections. Nat Med. 2020 Aug;26(8):1200–4.

11. Prévost J, Gasser R, Beaudoin-Bussières G, Richard J, Duerr R, Laumaea A, et al. Cross-sectional evaluation of humoral responses against SARS-CoV-2 Spike [Internet]. Microbiology; 2020 Jun [cited 2021 Dec 8]. Available from: http://biorxiv.org/lookup/doi/10.1101/2020.06.08.140244

12. Bates D, Mächler M, Bolker B, Walker S. Fitting Linear Mixed-Effects Models Using lme4. J Stat Soft [Internet]. 2015 [cited 2021 Dec 8];67(1). Available from: http://www.jstatsoft.org/v67/i01/

13. Zhang K, Ma Z-G, Yang L, Kang W, Yin Y, Lau JY-N. Significant reduction of humoral response to SARS-CoV-2 4 months after the diagnosis of COVID-19. Precision Clinical Medicine. 2021 Apr 3;4(1):73–6.

14. Seow J, Graham C, Merrick B, Acors S, Pickering S, Steel KJA, et al. Longitudinal observation and decline of neutralizing antibody responses in the three months following SARS-CoV-2 infection in humans. Nat Microbiol. 2020 Dec;5(12):1598–607.

15. Iyer AS, Jones FK, Nodoushani A, Kelly M, Becker M, Slater D, et al. Dynamics and significance of the antibody response to SARS-CoV-2 infection [Internet]. Infectious Diseases (except HIV/AIDS); 2020 Jul [cited 2021 Dec 8]. Available from: http://medrxiv.org/lookup/doi/10.1101/2020.07.18.20155374

16. Dan JM, Mateus J, Kato Y, Hastie KM, Yu ED, Faliti CE, et al. Immunological memory to SARS-CoV-2 assessed for up to 8 months after infection. Science. 2021 Feb 5;371(6529):eabf4063.

17. Wu J, Liang B, Chen C, Wang H, Fang Y, Shen S, et al. SARS-CoV-2 infection induces sustained humoral immune responses in convalescent patients following symptomatic COVID-19. Nat Commun. 2021 Dec;12(1):1813.

