## Supplemental File for "A novel assessment method for COVID-19 humoral immunity duration using serial measurements in naturally infected and vaccinated subjects"

### Supplement

**Table S1.** Individual results for the 20 naturally infected subjects. Dilution Factors corresponding to 50% reactivity for the first sample is presented for the 6 different antibodies. First and Last sample* indicates the time lapse in days since first symptoms of infection. N = number of samples for each subject. "≤ 6" means that the DF50-value was estimated below limit of quantification. Half-life are obtained from the linear regression model of ln(DF50) against time, in case the model returned a negative slope. Half-life greater than last sample time or when the model returned zero or positive slopes were annotated as > last sample day. Halftimes were indicated as Non-Estimable (NE) when slopes could not be estimated due to low first sample DF50 values (≤50).

| | | | | | DF50 of first sample | | | | | | halftime (days) | | | | | |
|---|---|---|---|---|---|---|---|---|---|---|---|---|---|---|---|---|
| Patient | N | Sex | First Sample* | last sample* | MP1 | NP1 | NP2 | RBD | S1 | S2 | MP1 | NP1 | NP2 | RBD | S1 | S2 |
| 1 | 5 | F | 32 | 267 | 16 | 680 | 38 | 1102 | 376 | 81 | >267 | 45 | NE | 106 | 119 | >267 |
| 2 | 5 | M | 16 | 360 | 110 | 273 | 435 | 706 | 743 | 112 | 91 | 82 | 66 | 196 | 124 | 240 |
| 3 | 5 | F | 26 | 358 | 7 | 859 | 27 | 3620 | 1524 | 434 | NE | 77 | NE | 133 | 115 | 218 |
| 4 | 4 | M | 33 | 265 | 6 | 294 | 52 | 11170 | 2325 | 323 | NE | 62 | 80 | 103 | 91 | >265 |
| 5 | 2 | M | 38 | 127 | ≤6 | 1086 | 56 | 1346 | 433 | 518 | NE | 42 | 32 | 29 | 34 | 31 |
| 6 | 6 | M | 29 | 270 | 25 | 7754 | 80 | 13376 | 5576 | 458 | NE | 25 | NE | 43 | 37 | 154 |
| 7 | 2 | M | 34 | 127 | 17 | 394 | 255 | 1876 | 489 | 5365 | NE | 16 | 17 | 11 | 15 | 10 |
| 8 | 7 | F | 26 | 270 | 7 | 44 | 10 | 229 | 129 | 93 | NE | NE | NE | 117 | 137 | 187 |
| 9 | 4 | F | 41 | 267 | 65 | 353 | 25 | 1759 | 549 | 1299 | 75 | >267 | NE | 927 | >267 | 194 |
| 10 | 6 | M | 37 | 359 | ≤6 | 88 | 137 | 5781 | 897 | 610 | NE | 74 | 90 | 67 | 129 | 111 |
| 11 | 5 | M | 38 | 358 | 10 | 1387 | 53 | 6667 | 3096 | 1203 | NE | 60 | 108 | 120 | 123 | 142 |
| 12 | 4 | F | 34 | 267 | 26 | 87 | 7 | 261 | 77 | 284 | 226 | 86 | NE | 170 | 256 | 113 |
| 13 | 5 | F | 24 | 268 | ≤6 | 536 | 18 | 566 | 262 | 53 | NE | 45 | NE | >268 | >268 | >268 |
| 14 | 6 | M | 19 | 270 | ≤6 | 1794 | 34 | 768 | 607 | 239 | NE | 37 | NE | 87 | 91 | 87 |
| 15 | 5 | M | 19 | 267 | 55 | 53 | ≤6 | 255 | 55 | 39 | 169 | 75 | NE | 164 | 230 | NE |
| 16 | 5 | F | 19 | 267 | 11 | 234 | 23 | 474 | 216 | 90 | NE | 148 | NE | >267 | 223 | >267 |
| 17 | 6 | M | 19 | 356 | 125 | 172 | ≤6 | 2240 | 941 | 213 | 74 | 66 | NE | 126 | 136 | 186 |
| 18 | 6 | M | 16 | 265 | ≤6 | 149 | 49 | 366 | 124 | 30 | NE | 60 | 101 | 120 | 153 | NE |
| 19 | 9 | F | 33 | 357 | 12 | 324 | ≤6 | 508 | 407 | 51 | NE | 118 | NE | 168 | 130 | >357 |
| 20 | 4 | M | 23 | 268 | 16 | 54 | ≤6 | 331 | 230 | 1637 | NE | 155 | NE | 119 | 103 | 76 |
| **Median** | **5** | | **29** | **268** | **11** | **309** | **31** | **935** | **461** | **262** | **-** | **66** | **-** | **120** | **127** | **187** |

**Sigmoidal curve fitting**

We used two different approaches to obtain DF50-values, but they are both based on the general form of the sigmoidal dose-response curve can be written as follows:

$$Y=Bottom + (Top-Bottom)/(1+2^{(Log_2 DF50-Log_2 DF)*Hillslope})$$

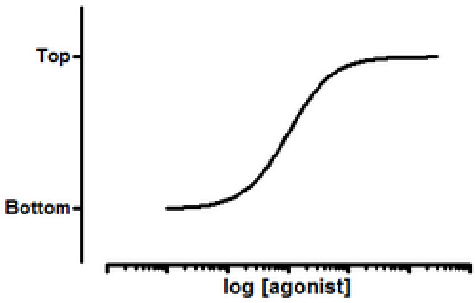


We simplified this 4-parameter curve to a 2-parameter sigmoidal function by fixing top and bottom in the above equation.

By using the Positive Control (PC) to normalize the reactivity of every biomarker reactivity, and present the reactivity as a percentage, we could set the top = 100 and bottom = 0; so we introduced the following constraints:

- Top = 100
- Bottom = 0

This reduces the model to

$$Y = 100 /(1+2^{(Log_2 DF50-Log_2 DF)*Hillslope})$$

Note that we use $\log_2$ because the dilutions are in a series of ½ but it does not really matter which base is used for the logarithm.

<u>Non-linear mixed modelling approach</u>

As explained in the main manuscript, non-linear mixed modelling of the above equation on the total collection of the samples was applied, in other words the hillslopes (per marker) are fitted on the combination of all 396 samples. At the same time, this is an advantage and disadvantage of the nonlinear mixed model. The advantage is robustness, with specific weighing of the dilutions, but the disadvantage is that it requires a 'collection' of samples, in other words, it cannot be used for one single sample.

Alternative individual modelling approach

This method is based on fitting the 2-parameter model for each dilution series separately, resulting in different hillslopes and DF50-values for each dilution series. This can be realized by converting the above non-linear 2-parameter model to a linear equation.

With some algebra, we have

$$(100 - Y) / Y = 2^{(Log_2 DF50 - Log_2 DF)*Hillslope}$$

Taking the $\log_2$ of both sides, we have

$$Log_2 [ (100 - Y) / Y] = (Log_2 DF50 - Log_2 DF)*Hillslope$$

Or

$$Log_2 [ (100 - Y) / Y] = Hillslope * Log_2 DF50 - Hillslope * Log_2 DF$$

Taking $Y' = Log_2 [ (100 - Y) / Y]$ and $X = Log_2 DF$, the expression above reduces to a linear relationship with slope = - Hillslope and intercept = Hillslope * $Log_2$DF50.

Therefore, by regressing Y' against X we are able to determine DF50 and the Hillslope.

Note that the above transformation is not possible in case Y = 100 or Y = 0. In these cases, we set Y = 99 and Y = 1 to avoid errors in the calculation. Special care should be taken when the reactivity (Y) is noise only, as in these cases the individual approach may return a DF50-value which is not reflecting the true sigmoidal behavior.

**Time-evolution of the DF50-reactivity**

We assumed an exponential decay of the biomarker reactivity with time, based on the following mathematical formula:

$$DF50 = A \exp(-t/\tau)$$

This equation can be transformed to a linear equation, by taking the natural logarithm of both sides:

$$\ln(DF50) = \ln(A) - t/\tau$$

Time t can be obtained as:

$$t = \tau \times (\ln(A) - \ln(DF50)] = \tau \times \ln[\frac{A}{DF50}]$$

The half-life $T_{50}$ can be defined as the time required to set DF50 = A/2, the time $T_{90}$ is obtained by setting DF50 = A/10. Therefore, the characteristic times are defined as

$$T_{50} = \tau \times \ln(2) \qquad \text{(half-life)}$$

$$T_{90} = \tau \times \ln(10))$$

As an example, we present the DF50-values of the naturally infected patient #1 in figure S1. The DF50-values were fitted against time (in days) resulting in $\tau$ = 1/0.006464 = 154.7. Consequently T50 = 154.7 x ln(2) = 107 days and T90 = 154.7 x ln(10) = 356 days. According to this model, 90% of the antibodies disappeared within 356 days after the infection.

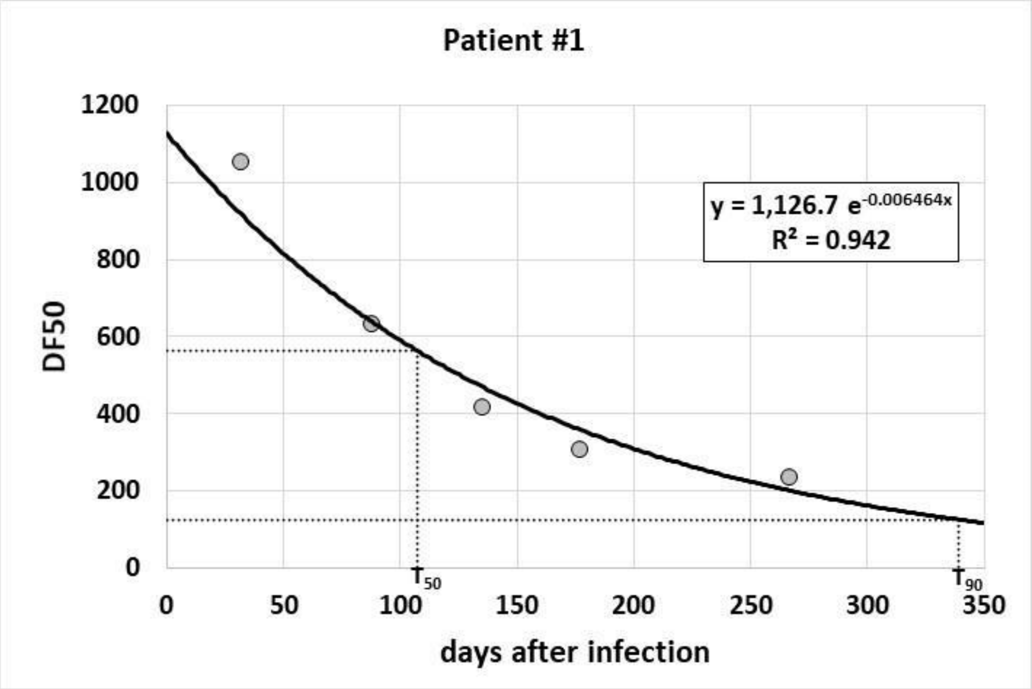


Figure S1. DF50 decay over time for the naturally infected patient #1
